# Prevalence and Factors Associated with Chronic Kidney Disease among Patients Admitted to Medical Ward in a Tertiary Hospital, Northern Ethiopia, a Cross Sectional Study

**DOI:** 10.1101/2022.04.04.22273431

**Authors:** Meskelu Kidu Weldetensae, Migbnesh Geberemedhin, Measho Gebreselassie, Ephrem Berhe

**Affiliations:** Nephrology Unit, Department of Internal Medicine, School of Medicine, College of Health Sciences, Mekelle University, Tigray, Ethiopia; Department of Health System, School of Public Health, College of Health Sciences, Mekelle University, Tigray, Ethiopia

**Keywords:** Chronic Kidney Disease, Hypertension, Nephrotoxic Agents, Recurrent Urinary Tract Infection

## Abstract

**Introduction:** Chronic Kidney Disease (CKD) is being recognized as a global public health problem. CKD is a major non-communicable disease with the global prevalence varying between 10.5% and 13.1%. Diabetes and hypertension appear to be the leading causes of CKD and End Stage Renal Disease worldwide. The aim of this study is to determine the prevalence of CKD and its associated factors among patients admitted to medical ward in a tertiary hospital, Northern Ethiopia.

**Methodology:** An institution based cross-sectional study was undertaken using systematic random sampling technique to select study participants. Sample sizes of 450 patients were included in the study. Data was collected using a pre-tested semi-structured questionnaire designed to meet the study objective. The data collection period was from October 20, 2017 to March 20, 2018 G.C. Data was analyzed using SPSS version 21.The odds ratio with their 95% confidence interval and P value were calculated. Statistical significance was declared if P value < 0.05.

**Result:** Of the 450 patients, 260(57.8%) were males. More than half (54.2%) were between ages of 25 to 40 years. The overall prevalence of CKD among patients admitted to medical ward was 17.3% (95% CI 13 - 29.9) and 14.4% (95 % CI 6.2 – 12.3) by Cockcroft Gault and MDRD equations respectively. Prevalence of stage 5 CKD was 61.5% by Cockcroft Gault equation. Hypertension AOR 3(95%CI 1.28, 4.1), history of recurrent urinary tract infection AOR 3.5 (95% CI 1.1, 7.3) and history of using nephrotoxic drugs AOR 3.4 (95% CI 2, 9.3) were significantly associated with CKD.

**Conclusion:** The prevalence of CKD among adult patients admitted to medical ward in tertiary hospital, Northern Ethiopia was high and majority of patients with CKD were stage 5. Hypertension, use of nephrotoxic agents and recurrent urinary tract infections were significantly associated with CKD.

## Background

Chronic Kidney Disease (CKD) is defined as the presence of markers of kidney damage for ≥ 3 months. These markers include structural or functional abnormalities of the kidney, pathological abnormalities or other kidney damages, including abnormalities in the composition of blood or urine, abnormalities in imaging tests, presence of Glomerular Filtration Rate(GFR)< 60 mL/min/1.73 m2(1). The presence of kidney disease is usually evaluated by GFR, an important marker of kidney function, and using indicators of kidney damage like proteinuria, abnormal urinary sediment, abnormal imaging studies and presence of kidney transplant(2).

CKD is a continually growing problem worldwide with estimated prevalence of ranging 10.5- 13.1 % and has a risk-multiplier effect on major non-communicable diseases, including cardiovascular diseases. Because of this CKD has become a major health problem globally, especially in developing countries with substantial effect in Sub-Saharan Africa(1,3).

The disease epidemiology is markedly different from other regions. Although middle-aged and elderly populations are predominantly affected in developed countries; in sub-Saharan Africa, CKD mainly affects economically productive young society between the ages of 20 to 50 years.

Furthermore, it is may not be associated with major risk factors like Hypertension, diabetes and smoking(4,5).

CKD is classified into 5 stages, as follows(6)

Stage 1: Proteinuria ≥30 mg/dl and eGFR>90 ml/min/ 1.73 m2

Stage 2: Proteinuria ≥30 mg/dl and eGFR 60 to 89 ml/ min/1.73 m2

Stage 3: eGFR of 30 to 59 ml/min/1.73 m2 with or without Proteinuria

Stage 4: eGFR of 15 to 29 ml/min/1.73 m2 with or without Proteinuria

Stage 5: <u>E</u>nd <u>S</u>tage <u>R</u>enal <u>D</u>isease (ESRD): eGFR<15 ml/min/1.73 m2 with / without Proteinuria

In addition to GFR, proteinuria has a significant role in assessing risk of CKD; especially important since patients with stage 1-2 CKD and proteinuria have worse outcomes than people with stage 3 and no proteinuria. Furthermore, the risk of development of ESRD and cardiovascular mortality is accurately predicted by proteinuria measurements than by GFR(7,8).

Chronic kidney disease is generally unrecognized in the early stages, as there are no particular symptoms. They could have nonspecific symptoms generally unwell feelings and decrease in appetite or indistinguishable symptoms like pallor, fatigue, gastrointestinal disorders like hiccups, nausea and vomiting or with symptoms of heart failure(9).

There is high prevalence of asymptomatic CKD among study populations(10,11). Usually, CKD is diagnosed during the evaluation of patients who have high blood pressure, diabetes and those who have family history. However, developing countries are highly affected with CKD that cannot be explained by diabetes or hypertension(3), around 43% of people with CKD do not have diabetes or hypertension(12). In most studies, epidemiological and clinical evidences have shown an increased risk for CKD among individuals with certain clinical and socio-demographic characteristics(8). However, in some places such as Sri Lanka and Nicaragua, the conventional risk factors were not associated with the disease prevalence(13,14).

## Methods

The study was conducted in a single center, institution based cross-sectional study in adult patients admitted in Ayder Hospital Medical ward which is located in Mekelle, capital of Tigray region northern part of Ethiopia, was conducted from October 20, 2017 - March 20, 2018 G.C.

### Sample Size and Sampling

#### Method Inclusion Criteria

Patients admitted to Ayder hospital medical wards that consented to participate in the study and aged ≥18 years were included.

#### Exclusion Criteria

Patients who had clear risk for acute kidney injury and/or diagnosed with acute kidney injury were excluded from the study.

#### Data Collection

All patients admitted in medical wards during the study periods had their charts examined, 572 patients were enrolled by using systematic random sampling technique to collect data through partially close-ended questionnaires, interview and document analysis. The study groups were sequentially included, after excluding those with feature of acute kidney injury such us recent rise in creatinine, sepsis, critically ill and shock patients as can be seen in Fig 1.

**Figure 1:**
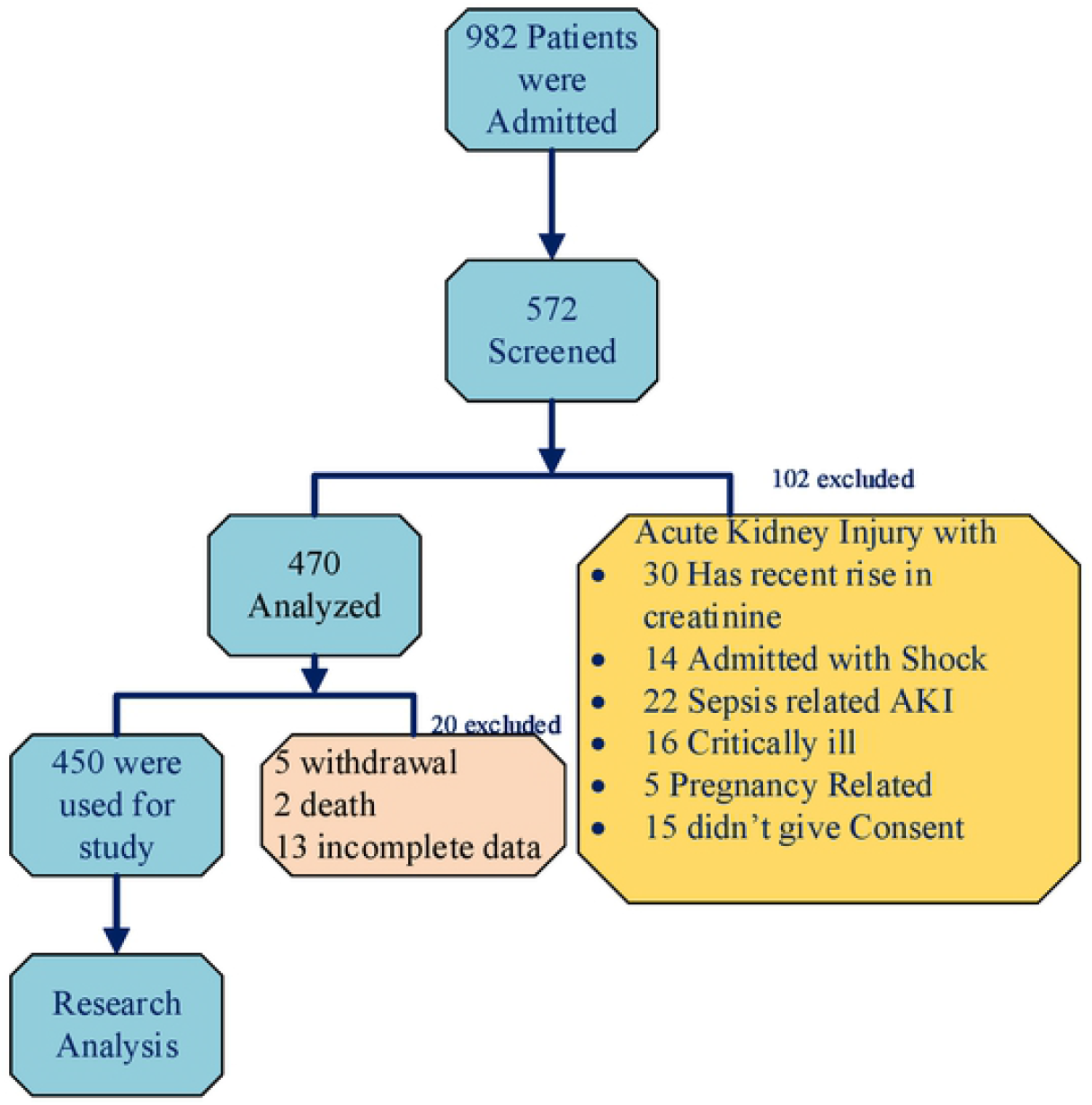
Data collection procedure flow chart

<u>B</u>lood <u>P</u>ressure (BP), Weight, height were measured by trained nurses. BMI was calculated as weight (in kilograms) divided by height in square meters. Those with newly detected high creatinine and having patient with difficulty to differentiate acute kidney injury and chronic kidney disease were advised to start further work up and appointed at least after 3 months for follow-up investigation and treatment at the hospital and it was revised from Smart Care using their identification number.

#### Data Analysis

The collected Data were coded and entered to Epi Data manager and cleaned, then exported to SPSS version 21 for analysis. Tabulation, cross tabulation, computation of frequencies and percentages were done on selected variables. Binary logistic regression analysis was used to check the association between single explanatory variable and dependent variable with odds ratio at 95% CI and statistical significance of 5%. Multivariable logistic regression was done to identify associated factors of CKD and to control for possible confounding effect. P-value < 0.05 was considered statistically significant.

## Results

Total numbers of patients admitted to medical ward within the study period were 982, of which 572 patients were screened. Then, the data of 450 patients was analyzed as depicted in Fig 1. The mean and SD of the age of the study participants was 45 ± 18 years and more than half (54.2%) of them were between 25 to 54 years of age. Table 1 shows, from the study, 260 (57.8 %) participants were males, with male to female ratio of 1.4:1.

**Table 1:** Socio demographic and clinical characteristics of patients admitted to medical ward (n=450)

| Variables | Frequency | Percentage |
| --- | --- | --- |
| **Gender** |  |  |
| Male | 260 | 57.8 |
| Female | 190 | 42.4 |
| **Age(years)** |  |  |
| 18-24 | 67 | 14.9 |
| **25-54** | 244 | 54.2 |
| **55-64** | 55 | 12.2 |
| **≥65** | 84 | 18.7 |
| **Residence** |  |  |
| Rural | 269 | 59.8 |
| Urban | 181 | 42.2 |
| Obesity (BMI >30kg/m<sup>2</sup>) | 6 | 1.3 |
| History of recurrent UTI | 130 | 28.9 |
| Body swelling | 163 | 36.2 |
| Family history of kidney disease | 10 | 2.2 |
| Diabetes mellitus | 41 | 9.1 |
| Duration of DM(<5 years) | 35 | 86.9 |
| Anemia | 214 | 47.6 |
| HBV infection | 16 | 3.6 |
| HCV infection | 14 | 3.1 |
| HIV infection | 127 | 28.2 |
| **Laboratory values** |  |  |
| eGFR | 91±52ml/min/1.73m<sup>2</sup> |  |
| Serum creatinine | 5.1 ±0.7 mg/dl |  |
| Urine dipstick proteinuria | 110 | 24.4 |
| **Abdominal ultrasound findings** |  |  |
| Small shrunken kidney | 60 | 13.3 |
| Enlarged kidney | 14 | 3.1 |
| Normal sized kidney | 344 | 76.4 |
| Poly cystic kidney | 18 | 4 |
| Obstructive Uropathy | 10 | 2.2 |
| Others* | 4 | 0.9 |

The prevalence of CKD was 17.3 % (CI 95% 13 - 29.9) and 14.4% (CI 95 % 6.2 –12.3) by Cockcroft Gault and MDRD equations respectively. Fig 2 demonstrates the prevalence of CKD classified by stages based on Cockcroft Gault equation, that is 48 patients (61.5 %) were stage 5, 12.8 % were stage 1, 7.8% were stage 3, 15.3% patients were in stage 4 CKD.

**Figure 2:**
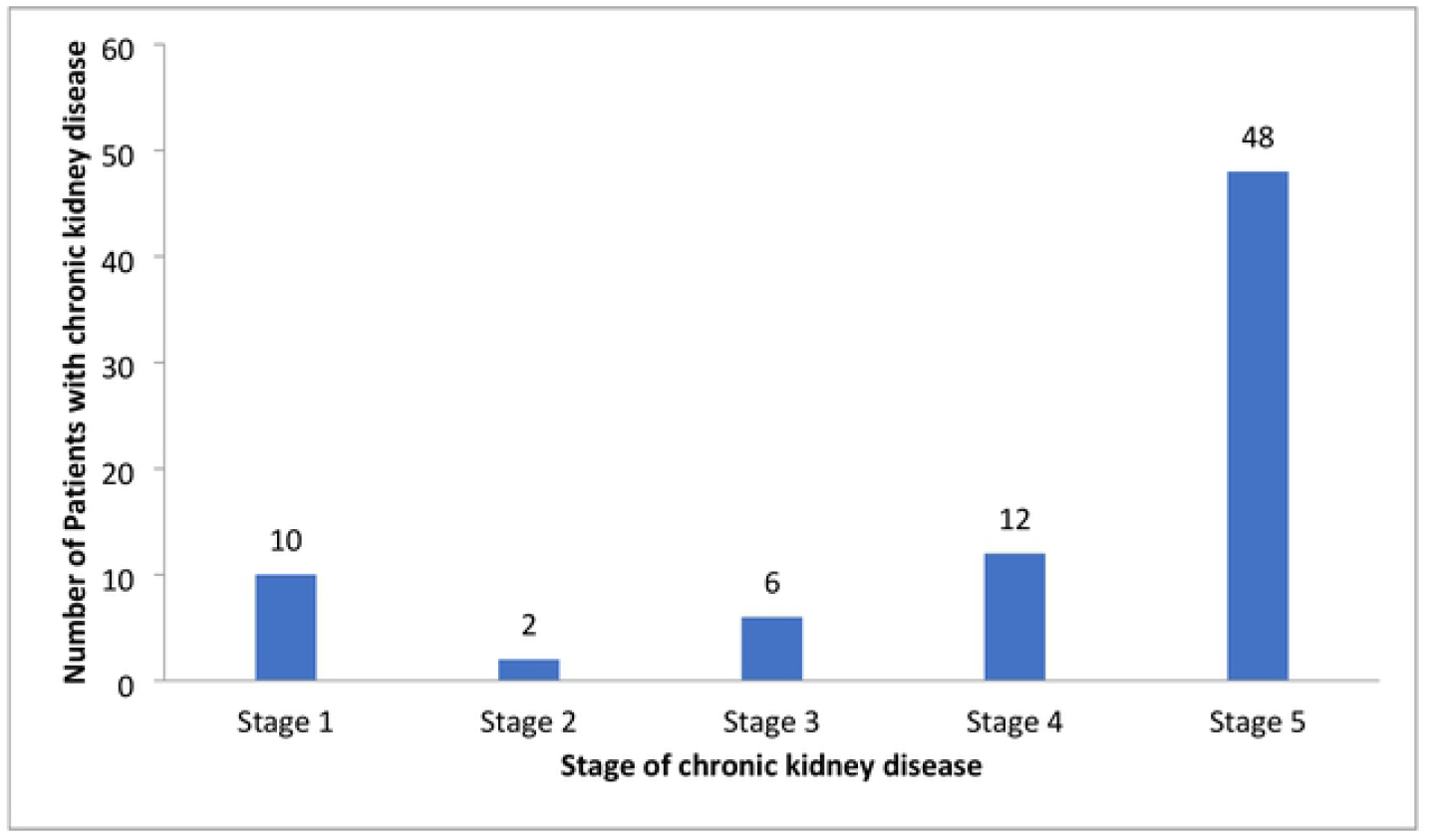
Prevalence of CKD classified by stages according Cockcroft Gault

In the multivariate analysis which can be shown in Table 2, patients with current hypertension were 3 times more likely to develop CKD than those with blood pressure < 140mmHg (AOR=3, 95% CI(1.28, 4.1), patients with history of recurrent UTI were 3.5 times more likely to develop CKD than patients without history (AOR=3.5, 95% CI (1.1, 7.3). Patients with history of using nephrotoxic drugs were 3.4 times at risk of developing CKD than those who had no similar history (AOR=3.4, 95% CI (2,9.3).

**Table 2:** Bivariate and multivariate analysis of factors associated with CKD among patients admitted to medical ward

| Variable | Response | CKD |  | COR (95% CI) | AOR(95%CI) | P-value |
| --- | --- | --- | --- | --- | --- | --- |
|  |  | Yes | No |  |  |  |
| Age | >65 Years | 14 | 70 | 1.1(0.5 ,1.9) |  |  |
|  | <65 years | 64 | 302 | 1 | 1 |  |
| Gender | Male | 52 | 208 | 0.63 (0.38, 1) |  |  |
|  | Female | 26 | 164 | 1 | 1 |  |
| Smoking status | Yes | 19 | 40 | 2.6 (1.4 ,4.9) | 1.8(0.83,4.2) | 0.129 |
|  | No | 59 | 332 | 1 | 1 |  |
| History of exposures to nephrotoxic drugs | Yes | 9 | 74 | 1.11(1.2 , 1.8) | 3.4 (2, 9.3) | **0.001** |
|  | No | 69 | 298 | 1 | 1 |  |
| HIV infection | Yes | 20 | 107 | 1.7 (0.6 , 2) | 0.58(0.4,1.4) | 0.80 |
|  | No | 58 | 265 | 1 | 1 |  |
| History of recurrent UTI | Yes | 53 | 77 | 8.1 (4.7, 13.9) | 3.5(1.1 ,7.3) | **0.01** |
|  | No | 25 | 295 | 1 | 1 |  |
| Family history of CKD | Yes | 2 | 3 | 1.3(0.05 ,1.8) | 1.2(1.0 , 6.3) | 0.161 |
|  | No | 76 | 368 | 1 | 1 |  |
| Other chronic medical illness | yes | 30 | 185 | 1.5(0.9 ,2) | 1.3(1.3 ,9.6) | 0.14 |
|  | no | 48 | 187 |  | 1 |  |
| BMI | >30 | 3 | 3 | 4.9 (0.9 ,20) | 2.1 (0.2 ,16) | 0.44 |
|  | <30 | 75 | 369 | 1 | 1 |  |
| Systolic BP | ≥140 | 47 | 59 | 8(4.7, 13.6) | 3(1.28 ,4.1) | **0.017** |
|  | <140 | 31 | 313 | 1 | 1 |  |
| Fasting blood sugar | ≥126 | 13 | 28 | 2.4(1.2 ,4.9) | 0.69(2.2,10.7) | 0.24 |
|  | <126 | 65 | 344 | 1 | 1 |  |

## Discussion

In this study majority of the patients were males accounting for about 57.8 % of the patients. More than half of patients were between 25-54 years of age. This is similar to a study done in Uganda where 51.3 % of patients were males and with median age of 39 years(15). Similarly according to the study in sub-Saharan Africa, CKD mainly affects economically productive young society between the ages of 20 and 50 years; and was not associated with major risk factors like <u>H</u>yper<u>t</u>ensio<u>n</u> (HTN), diabetes and smoking(16,17), this might be due to most nontraditional risk factors like infectious cause are highly prevalent.

Prevalence of CKD based on eGFR from this study was 17.3 % (95% CI 13 - 29.9) among hospitalized patients to medical ward. This is similar to the study done in Uganda that found 18 % among hospitalized patients(15). The participants in the Uganda study were critically ill and hence the slight increase in total number of renal dysfunction cases. In a few in-patient studies in Africa, renal dysfunction ranged from 17% to almost 30% in HIV positive patients(18). The difference is that the African studies recruited patients with surgical and obstetric conditions as well. In a population based cohort study in Germany in 2012 the prevalence of CKD in community dwelling elderly and study done in a rural region of Haiti from patients that visit the outpatient’s department showed prevalence of 24% and 27% respectively(19,20). This is slightly higher than our study that is because they might have included elderly patients and used ACR to calculate proteinuria. Furthermore, the study didn’t use ACR categories to diagnose CKD which can underestimate the prevalence. But, another study done in Switzerland and United States showed lower prevalence, that is, 13.8 and 10%, respectively(21,22). The reasons for the differences in the above mentioned studies and this study might be due to, the above results have conducted population based study with distributed risks and partly, might be due to our hospital being only referral hospital where patient’s with complication requiring renal replacement therapy are also admitted.

Majority of patients in this study present with stage 5 CKD (ESRD) in contrast to other studies where the prevalence of CKD stage 3 to 5 in the adult US population according to the third National Health and Nutrition Examination Survey (NHANES 3) was only 4.7%(22) and similarly in Swiss; stage 4 and 5 was only 4.5 %(19). The reason why patients present in ESRD in this study might be because of the absence of screening of patients with known risk factors for CKD. To some extent, it can also be due our hospital is the only center with RRT for patient presenting with complication. This is consistent with many of African countries study like, Uganda and Botswana(15,23).

Worldwide hypertension is recognized as one of the leading causes of CKD(24). This study revealed that patients with systolic hypertension were 3 times more likely to develop CKD than the non-hypertensive patients. Studies done in Uganda, Haiti and DRC also found correlation between hypertension and CKD(5,15,19). The use of nephrotoxic agents has been associated with CKD as demonstrated in Burundi(25). Similarly, in the present study the association of nephrotoxic drugs with CKD was 3.4 times than non-users. However, in Uganda, it was not found to be associated, this can be due to they have only assessed NSAID use unlike this study that include other nephrotoxic agent including contrast media(15).

While diabetes in most studies is mentioned as an independent predictor for CKD, in this study the prevalence of diabetes in the participants was 9.1 % (95 % CI 6.4-12.0); however in the multivariate regression model it was not significantly associated with CKD. A probable explanation for this might be the recent development of diabetes in the study participants. This is similar to study done in Haiti and Iran(19,26). Infectious disease are known risk factors of CKD, but HIV infection was not associated with CKD in this study which is inconsistent with many African studies like Zambia and Botswana(15,23) This can be explained by the high prevalence of HIV in non CKD patients leading to distribution of risk factors.

### Limitation of the Study

The study population consisted of patients admitted to medical wards, population of higher risk for CKD, and does not represent the general population. Urinary Albumin-Creatinine ratio was not assessed, hence there is a possible underestimation of the prevalence of patients with proteinuria in this study especially stage 1 and 2 CKD. As this is a cross-sectional study design makes it may be impossible to infer a causal relationship between CKD and the risk factors.

## Conclusion

In conclusion, we found a high prevalence of CKD among adult patient admitted to medical ward of Ayder Hospital and majority of them were CKD stage 5 (ESRD). In this study significant association was demonstrated between CKD and elevated systolic blood pressure, history of using nephrotoxic drugs and history of recurrent urinary tract infection.

### Recommendation

The study recommends further investigations encompassing a large number of population to find out the point prevalence of CKD and its associated factors so that a preventive strategy especially for the preventable associated factors or for an entire defensive framework could be implemented or designed to reduce the expansion of the disease at community level.

These data have to be noted by the health authorities in the country to integrate CKD in public health planning with the aim to develop adapted preventive strategies and give health education to the community about the importance of screening modalities in high risk patients and especially regarding the use of nephrotoxic drugs.

## Data Availability

The dataset generated and analyzed during this study is available and can be shared from the corresponding author upon reasonable request.

## Ethical Approval and Consent to Participant

Ethical clearance waived from Mekelle University, College of Health Sciences, and Institutional Research Review Board (IRRB). Permission received from the medical director of the hospital before the commencement of the study and informed written consent was also obtained from all eligible participants. In addition consent of participants who were unable to read and write was obtained from their legal representatives. The study adhered to relevant guidelines and regulations.

## Consent for Publication

Not applicable for this manuscript

## Availability of Data and Materials

The data set generated and analyzed during this study is available and can be shared from the corresponding author upon on reasonable request.

## Computing Interest

The authors declare that they have no computing interest.

## Funding

The cost of this research was covered by Mekelle University. The funder had no role in study design, data collection and analysis, decision to publish, or preparation of the manuscript

## Author’s Contribution

**Conceptualization:** Meskelu Kidu Weldetensae, Migbnesh Geberemedhin, Measho Gebreselassie

**Data curation:** Meskelu Kidu Weldetensae, Migbnesh Geberemedhin,

**Formal analysis:** Meskelu Kidu Weldetensae, Migbnesh Geberemedhin, Measho Gebreselassie

**Funding acquisition:** Meskelu Kidu Weldetensae, Migbnesh Geberemedhin, Measho Gebreselassie

**Investigation:** Meskelu Kidu Weldetensae, Migbnesh Geberemedhin, Measho Gebreselassie

**Methodology:** Meskelu Kidu Weldetensae, Migbnesh Geberemedhin, Measho Gebreselassie

**Project Administration:** Meskelu Kidu Weldetensae

**Resources:** Meskelu Kidu Weldetensae

**Software:** Meskelu Kidu Weldetensae, Ephrem Berhe,

**Supervision:** Meskelu Kidu Weldetensae, Migbnesh Geberemedhin, Ephrem Berhe

**Validation:** Meskelu Kidu Weldetensae, Migbnesh Geberemedhin, Measho Gebreselassie, Ephrem Berhe

**Visualization:** Meskelu Kidu Weldetensae, Migbnesh Geberemedhin, Measho Gebreselassie, Ephrem Berhe

**Writing – Original Draft:** Meskelu Kidu Weldetensae

**Writing – Review & Editing:** Meskelu Kidu Weldetensae, Migbnesh Geberemedhin, Measho Gebreselassie,

## Acknowledgment

We are grateful to acknowledge data collectors and administrations of Ayder Comprehensive Specialized Hospital, Mekelle University. We also thank for the study participants.

## Notes

### Competing Interest Statement

The authors have declared no competing interest.

### Funding Statement

The cost of this research was covered by Mekelle University (MU). MU had no role in study design, data collection and analysis, decesion to publish, or preparation of the manuscript.

### Author Declarations

Ethical evidence waived from Mekelle University, College of Health Sciences, and Institutional Research Review Board (IRRB). Permission received from the medical director of the hospital before the commencement of the study and informed written consent was also obtained from all eligible participants. In addition consent of participants who were unable to read and write was obtained from their legal representatives. The study adhered to relevant guidelines and regulations.

